# Modelling mass asymptomatic testing strategies for early containment of infectious disease outbreaks in prisons

**DOI:** 10.64898/2026.03.13.26348273

**Authors:** Joseph Brooks, Francesca Scarabel, Jingsi Xu, Pauline Bakker, Ian Hall, James Adamson, Rupert Bailie, Rachel Campbell, Nicola Dennis, Lianne Straus, Steve Willner, Jonathan Van Der Veen, Chantal Edge, Tom Fowler, Lorenzo Pellis

## Abstract

**Objective:** Investigate a strategy of mass asymptomatic testing and isolation (“pulse testing”) aimed at early containment of outbreaks in prisons in comparison to or combination with a symptom-based isolation strategy.

**Methods:** Simulations using an individual-based time-since-infection model were run under different pathogen and intervention strategy scenarios. Measured outcomes were the proportion of outbreaks contained and number of individuals isolated.

**Results:** For *R*_0_ = 2, 25% probability of being asymptomatic (*p*_*a*_ = 0.25), a COVID-19-like infection dynamics and perfect adherence, one pulse test contained approximately 20% of outbreaks, and three tests up to 50%. With no asymptomatic cases, three tests performed similarly to isolating cases one day after symptoms (≈ 55% outbreaks contained), but symptom-based isolation degraded significantly faster than pulse testing with increasing *p*_*a*_. With perfect adherence, combining both interventions contained between ≈ 25% (*R*_0_ = 3, *p*_*a*_ = 0.5) and *>* 90% (*R*_0_ = 1.5, *p*_*a*_ = 0) of outbreaks. Across all scenarios, pulse testing isolated substantially fewer individuals than symptom-based isolation, e.g. ≈ 5% versus ≈ 30% for *R*_0_ = 2 and *p*_*a*_ = 0.25.

**Conclusion:** If implemented promptly upon outbreak declaration and with high adherence, pulse testing may stop outbreaks early, substantially reducing the number of isolations and mitigating the impact on prison regime and resident/staff wellbeing. However, for large *R*_0_ or delayed implementation, effectiveness drops rapidly.

## Introduction

The increased vulnerability of prisons to outbreaks of infectious diseases such as influenza and tuberculosis is well recognised [1–4], and during the COVID-19 pandemic outbreaks in prisons were reported worldwide [5, 6]. Upon the introduction of an infectious pathogen into an incarcerated population, the close proximity of prison residents to each other (perhaps due to overcrowding), sharing of facilities and poor air ventilation can lead to a rapid spread of infection. Individuals in contact with the criminal justice system also face huge health inequalities, experiencing much higher rates of mortality, smoking [7], chronic respiratory disease (including asthma), immunosuppression (for example, due to HIV infection), and other chronic illnesses (such as cardiovascular disease, diabetes, or liver disease) [8, 9], all of which increase the probability of severe outcomes from infection. This combination of a clinically vulnerable population and a closed environment makes explosive outbreaks of respiratory viruses likely and the impacts potentially severe [10]. These issues are exacerbated by the substantial costs and logistical challenges associated with managing regime operation during an outbreak in a prison estate, and there is a high potential for prison healthcare systems to be overwhelmed or constrained in their service delivery. For these reasons, it is important to study in advance how outbreaks of novel pathogens may be managed and investigate interventions which would not be considered feasible or cost-effective in other settings.

In the case of an outbreak of an infectious pathogen, non-pharmaceutical interventions (NPIs) may be deployed to reduce the rate of transmission between individuals, especially if pharmaceutical interventions such as vaccines or antivirals are not available. Interventions such as quarantining, screening, contact tracing and epidemiological surveillance have been used to control a variety of pathogens in the past [3]. During the COVID-19 pandemic, NPIs were shown to be effective in preventing infection [11] and potentially necessary to prevent prisons becoming amplifiers for infection in the community [6]. In the UK, prisons implemented a number of control measures such as quarantining on entry to prevent introduction or re-introduction [12].

However, some NPIs are also known to have detrimental effects on an individual’s well-being – effects that are more acutely felt by vulnerable individuals such as those in prison. Mental disorders are over-represented in the incarcerated population [13], with high rates of mental morbidities such as major depression (6-month prevalence of 11.4(9.9, 12.8)%) post-traumatic stress disorder (9.8(6.8, 13.2)%) and psychotic illness (3.7(3.2, 4.1)%) [14]. NPIs have been shown to disrupt mental health services in prisons [15] and further limitations on residents’ freedoms have serious impact on their mental health [16, 17]. For these reasons, particularly in this context, it is important to explore interventions that are effective in reducing the number of infections whilst minimising the potential for detrimental effects on wellbeing.

Mathematical models of infectious diseases have a long history of use in appraising NPIs in carceral settings [18]. During the COVID-19 pandemic, mathematical modelling has been used to assess several NPIs adopted in prisons, including cohort isolation, depopulation, single celling and asymptomatic testing [19–25]. Here we used a stochastic, individual-based, time-since-infection model to investigate the efficacy of two NPIs in containing an infectious disease outbreak in a prison.

First, we investigated the potential of a programme of mass asymptomatic testing early on in an outbreak, with subsequent isolation of individuals who test positive, to prevent the outbreak from becoming large and avoid longer-term interventions. In particular, we considered a protocol of testing all individuals in an affected population simultaneously, regardless of symptom status, using one or more “pulse” tests. We call this intervention “pulse testing and isolation” (PTI). Mass testing strategies have previously been used in English prisons [26] and the various potential uses of mass testing are central to future pandemic planning [27], particularly now that advances in diagnostic technology have reduced costs, facilitated self-testing (thus improving acceptability and uptake [28]) and opened up the option of testing multiple pathogens simultaneously [29].

Like other mass testing strategies, PTI has the potential to identify asymptomatic and presymptomatic cases, which posed a large problem during the COVID-19 pandemic [30, 31]. However, the intervention we discuss here differs from previously considered ones since it is implemented early on, shortly after outbreak declaration (in this work, following the detection of two epidemiologically-linked symptomatic cases), with the aims of interrupting the outbreak and preventing a large number of cases. Although it may appear an extreme intervention, if successful PTI can stop an outbreak early on, saving on resources and mitigating the impact on wellbeing in the prison. To reduce the logistical resources, PTI can be effective even if applied to a single wing or floor, as long as the tested population is sufficiently isolated from the rest of the prison. In our simulations, we considered a population of 100 susceptible individuals, approximately the scale of a single wing of a typical prison estate in England.

In this work we model the test as administered with a Lateral-Flow Device (LFD), in the sense that we assume instantaneous results (LFD results are typically available within 30 minutes), but imperfect sensitivity (maximum 95% at symptom onset). If needed, the approach could easily be modified to model a polymerase chain reaction (PCR) test (e.g. assuming nearly perfect sensitivity and at least a day delay to results), but LFDs are significantly cheaper, logistically easier (e.g. quick decision about isolation following testing) and, given they measure the presence of antigen, their positivity may be assumed to correlate better to an individual’s transmissibility than PCR tests [32].

We further compared PTI to a strategy of symptom-based isolation (SI) in which individuals who develop symptoms are subsequently isolated. Differently from PTI, SI not only reduces the probability of a large outbreak occurring, but also reduces the final size of all outbreaks. SI also differs from PTI since it requires isolating positive cases throughout the whole outbreak, whereas isolation from PTI is restricted to the few individuals identified early on. However, the prison system can implement SI even with no testing capacity, minimal clinical staff and little public health support. For this reason, we also explore the impact of implementing both interventions at the same time, viewing PTI as a potential additional intervention to SI, rather than exclusively as an alternative.

In this work we assume positive cases are isolated by themselves, to avoid any further transmission. This assumption presupposes that enough separate isolation facilities can be made available during an outbreak, which might not be feasible, and particularly so for the larger numbers occurring using an SI strategy. Prisons could alternatively opt for isolating positive cases in their cells, which in England are generally shared between two prisoners, though we have not modelled the impact of cell isolation here. We investigated and compared the effectiveness of PTI, SI and their combination under several scenarios, including different pathogen characteristics (transmissibility, infection period and probability of being asymptomatic), implementation strategies (delay from outbreak declaration to pulse test and number of pulse tests), and adherence of the individuals to the intervention. Since the effectiveness of mass testing is strongly related to the sensitivity of the test, and the effectiveness of isolation to the temporal profile of the probability of transmission, we considered a model in which both the test positivity profile and the individual infectiousness are correlated via a single underlying viral load profile, which depends on the time elapsed since infection. Our scenarios focused on a rather fast-paced disease with a generation time of 5 days, with pre-symptomatic and asymptomatic transmission, and medium transmissibility, reminiscent of respiratory infections such as influenza and COVID-19.

## Materials and Methods

### Infection Model

In this paper we used a stochastic, time-since-infection model in which an individual’s viral load, *V* (*τ*), is a function of an individual’s time since infection in days, *τ*. Since our interest is not in the quantitative details of the viral load but rather in how it changes over time, we chose units of measure so that the viral load *V* (*τ*) is normalised and 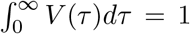. Specifically, we assumed that *V* (*τ*) = *g*(*τ* | *µ, σ*), where *g*(·|*µ, σ*) is the density function of a Gaussian distribution with mean *µ* and standard deviation *σ*. To restrict the distribution to only positive real times, the Gaussian curve was truncated at 0 and renormalised appropriately (though the rescaling correction is minimal in all profiles considered, as the mass truncated is never larger than 1%). We assumed *µ* = 5 days (the generation time) and we explored different values of *σ*, namely *σ* = 0.5, 1 and 2. These different cases are also referred to by the profile’s infection period, defined as the length of the period over which 95% of the viral load occurs (we use the word “infection” rather than “infectious/infectivity” to remind this period describes not only the period of high infectiousness, but also of high test positivity). The infection period is centred on the peak and its duration is given by the formula

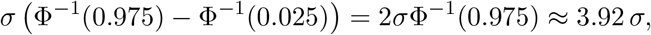

where Φ is the cumulative density function of the standard Gaussian distribution and Φ^−1^ its inverse. For *σ* = 0.5, 1 and 2, the lengths of the infection periods are approximately 2, 4 and 8 days, respectively. The viral load profile for *σ* = 2 matches closely to the one fitted by Ferretti et al. [33] to COVID-19 data, so we will refer to this as a COVID-19-like profile. We assumed that each infected individual makes infectious contacts at a rate proportional to their viral load: *λ*(*τ*) = *R*_0_*V* (*τ*), where *R*_0_ is the basic reproduction number. Individuals do not explicitly recover, but, since lim_*τ*→∞_ *λ*(*τ*) = 0, an infected individual’s contribution to new cases eventually becomes negligible.

Infected individuals can be either symptomatic or asymptomatic, but this was assumed to have no effect on the individuals’ viral load, transmissibility and test positivity profiles, and only served to determine when pulse testing is initiated and when individuals are isolated under SI. Individuals are asymptomatic with probability *p*_*a*_ and symptomatic with probability 1 − *p*_*a*_. Symptomatic individuals were assumed to develop symptoms deterministically at the peak of their viral load, exactly 5 days after infection.

### Non-pharmaceutical Interventions

We investigated and compared two types of interventions: pulse testing and isolation (PTI) and symptom-based isolation (SI). PTI takes place shortly after the declaration of an outbreak. An outbreak is assumed to be declared once there have been two symptomatic cases cumulatively, in line with the “two or more epidemiologically linked cases within five days” rationale currently used in prisons [8]. Since we are modelling an outbreak caused by one introduction of a single infection, any two cases in our model are epidemiologically linked by definition. After an outbreak is declared, pulse tests are implemented after a predetermined length of time, denoted by *T*_*k*_ for the *k*^th^ test. We considered intervention strategies that involve 1, 2 or 3 pulse tests. When a pulse test takes place, the whole population, regardless of infection and symptom status, is tested. Any individuals who test positive are isolated immediately (we assume the use of LFD, i.e. rapid results, although a delay can be added if needed) and cease to spread infection for the rest of the outbreak. For an infected individual, the probability of testing positive was also assumed to be proportional to their viral load according to the relation 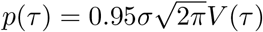. In particular, if a test is timed exactly with the peak of infection, there is a 95% chance that the test will result positive, a value also inspired by the sensitivity of LFDs for SARS-CoV-2 [34].

SI was modelled by isolating any symptomatic individuals after a fixed delay from the onset of their symptoms, until they are no longer infectious. Throughout this paper the delay was taken to be 1 day. With the SI intervention, isolation of symptomatic individuals started from the very beginning of the outbreak, independent of outbreak declaration, and until the end of the simulation.

To model adherence to the interventions, we assumed that each individual would either adhere to all interventions, or never adhere to any. For example, when 2 pulse tests were performed, an individual who rejects the first test/isolation will also reject the second. The adherence status of each individual was sampled randomly, with probability of adherence denoted by *p*_*ad*_.

### Stochastic Simulations and Scenarios

Since the effectiveness of a pulse test depends heavily on the timing and the time since infection of all individuals at the time of the test, we were unable to make use of analytical results known about similar models [35, 36], and instead had to simulate the full outbreak dynamics. To explore a large range of scenarios, we limited the simulations to a relatively small population, with *n* = 100 initial susceptibles and one single infective (*m* = 1). This population size is roughly comparable with the size of a single prison wing. For numerical efficiency, we adopted the construction first suggested by Sellke [37], adapted to time-inhomogeneous rates. Details can be found in Appendix A.

We considered several scenarios of hypothetical pathogens which varied in: the probability of a case being asymptomatic, *p*_*a*_; the basic reproduction number, *R*_0_; and the standard deviation for the Gaussian viral load profile, *σ*.

We investigated the effectiveness of 1, 2 or 3 pulse tests at containing the outbreak, and looked at how delays in their implementation, measured in days since the outbreak declaration and denoted by *T*_1_, *T*_2_ and *T*_3_, respectively, impact the effectiveness. We also investigated how the effectiveness of interventions changed with different probabilities of adherence *p*_*ad*_. Since we used a stochastic model, we ran 4000 simulations for each parameter set.

We utilised the Sellke construction of the epidemic to employ a coupling between epidemics, allowing us to make direct counterfactual comparisons between our simulated scenarios. Before running simulations we generated independent Exponential(1) distributed thresholds, denoted 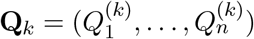 for *k* = 1, … 4000. These thresholds were then reused for each scenario. Each outbreak was conditioned on there being at least 5 individuals infected (5% of the susceptible population) under no interventions so as to only include outbreaks that did not die out quickly by chance. Whether an individual tests positive following a pulse test was coupled in a similar way to the infection events. Therefore, for each simulation with an intervention in place we could also simulate what would have happened with no intervention. This allowed us to use the following specific definition of containment: an outbreak is contained by an intervention if the final size is reduced to less than half of what it would have been without the intervention. We used the proportion of outbreaks contained by an intervention as a measure of its effectiveness. Since this is a binary measure (outbreaks are either contained or not) we construct binomial confidence intervals (CIs). Note that these only represent uncertainty from the stochasticity and are dependent on the number of simulations we run, i.e. they are not representative of uncertainty in any parameters.

## Results

### Effectiveness of Pulse Testing and Isolation

We first investigated the effectiveness of PTI in terms of percentage of outbreaks contained, focusing in particular on the differences between one single test or a combination of two or three tests in sequence.

The upper panels in Figure 1 show the rate of transmission (red, left axis) and test-positivity (blue, right axis) as function of the time since infection corresponding to each of the three different viral load profiles we considered. For each viral load profile, the bottom panels in Figure 1 show the percentage of simulated outbreaks that were contained using 1 single pulse test implemented after a delay *T*_1_ from outbreak declaration, specified by the horizontal axis. The colour and line styles correspond to different values of *R*_0_ and *p*_*a*_. The three panels show different trends. Panel F, corresponding to a long, COVID-19-like infection period, shows that, in general, the later a pulse test was, the less likely it was to contain an outbreak. For example, if *p*_*a*_ = 0 and *R*_0_ = 2 (Panel F, solid blue line), the percentage of outbreaks contained dropped from 24.8% if the test was immediately after the outbreak declaration (*T*_1_ = 0), down to 7.7% if the test was delayed by a week (*T*_1_ = 7). However, Panel D (narrow infection period) shows a clear peak in the percentage of outbreaks contained at *T*_1_ = 5, followed by a lower peak at *T*_1_ = 10. The intuitive explanation is that the narrow infection period leads to distinct, almost synchronised, generations of cases and, consequently, to a strong oscillatory pattern in the proportion of outbreaks contained, because a test performed when all cases in a generation are around the peak of their test-positivity profile will detect most of them, while a test performed in-between such peaks will likely miss them. A similar oscillatory pattern, although with a milder effect, is observed for a medium-length infection period (Panel E).

**Figure 1.**
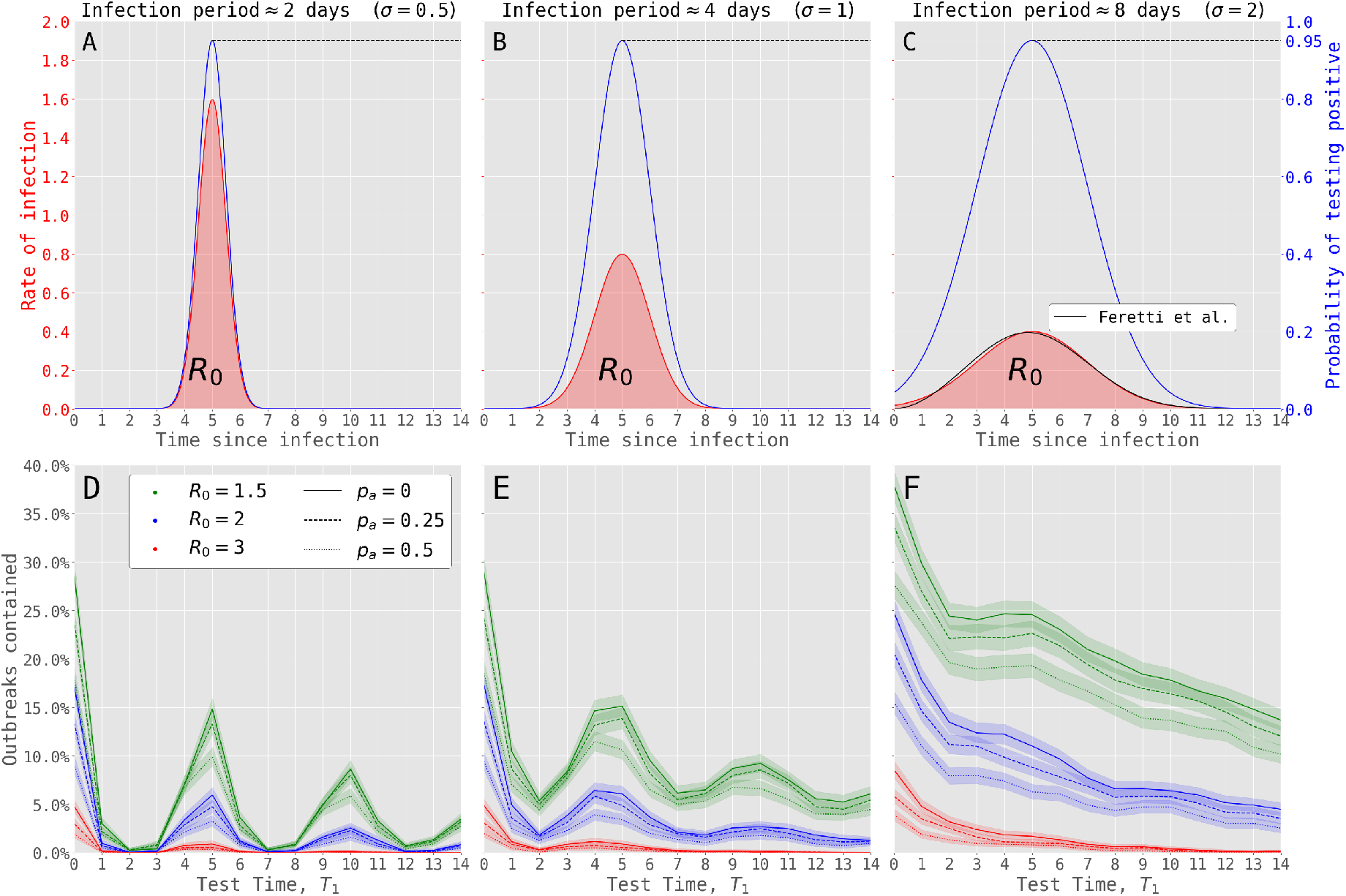
The three infectivity and test-positivity profiles are shown on the top row (panels A, B, and C). Each one is a scaled Gaussian curve with a different standard deviation (*σ*). The probability of testing positive is shown in blue and is chosen to peak at a 0.95. The rate of infection is shown in red and scaled so that the area under the curve is a chosen value of *R*_0_. In Panel C, the black curve shows the profile calculated by Ferretti et al. [33] from early COVID-19 data, which closely matches the profile with *σ* = 2. Below each profile is the corresponding percentage of outbreaks that are contained by a single pulse test depending on the time delay *T*_1_ from outbreak declaration (Panels D, E, and F). The colour and style of each line refer to the value of *R*_0_ and *p*_*a*_, respectively. Adherence is assumed to be perfect. Shaded areas show the 95% binomial CIs.

Figure 1 suggests that, despite perfect adherence, a single pulse test was unlikely to contain an outbreak in most of the scenarios we considered. Even in the most optimistic of these scenarios (*σ* = 2, *R*_0_ = 1.5, *p*_*a*_ = 0 and *T*_1_ = 0) only 37.9% of outbreaks are contained. In a less favourable scenario (*σ* = 2, *R*_0_ = 2, *p*_*a*_ = 0.25 and *T*_1_ = 0) this percentage drops to 20.6%. For this reason, in Figure 2 we explored the efficacy of implementing 1, 2 and 3 pulse tests in sequence, again in terms of percentage of outbreaks contained. Each subplot corresponds to a different infection profile (columns) and delay for the first pulse test (rows). The top row shows results for the first pulse test performed immediately following outbreak declaration (*T*_1_ = 0) and the bottom row shows results for a 1-day delay in the first test (*T*_1_ = 1). The timing of the second test (*T*_2_), where it applies, is determined by the *x*-axis. To reduce the complexity of the plot, the timing of the third pulse test, where it applies, was assumed to be 1 day after the second (*T*_3_ = *T*_2_ + 1). The thickness of the lines increases with the number of pulse tests and the colour determines the value of *R*_0_. In this figure, we assumed a 25% probability of cases being asymptomatic (*p*_*a*_ = 0.25).

**Figure 2.**
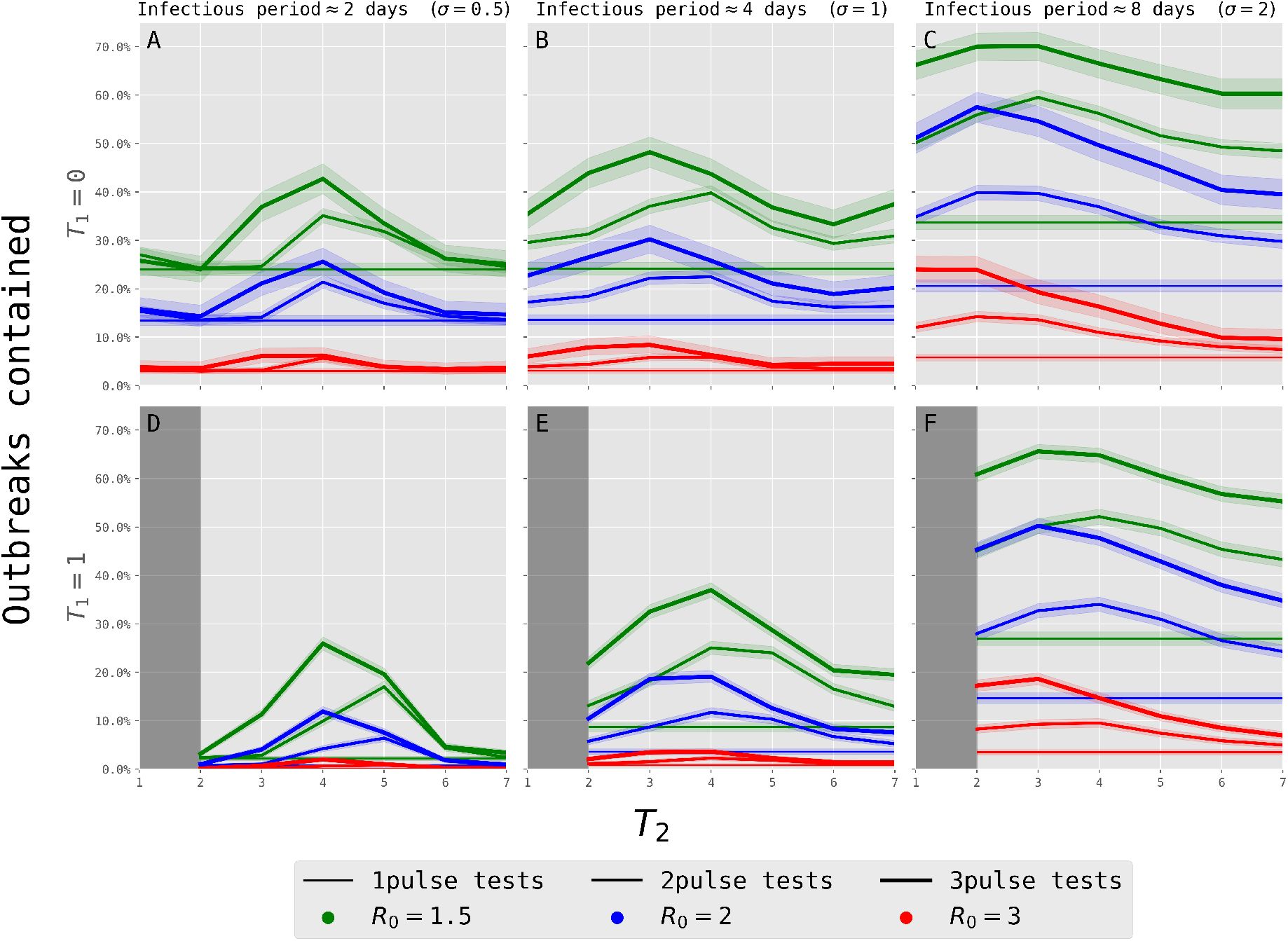
The percentage of outbreaks contained by 1, 2 and 3 pulse tests. The delay between the outbreak declaration and the first pulse test (*T*_1_) is determined by the row and the delay for the second test (*T*_2_), when it applies, is determined by the *x*-axis. The delay for the third test is 1 day after the second (*T*_3_ = *T*_2_ +1). We show results for *R*_0_ = 1.5 (green), 2 (blue) and 3 (red). Shaded areas show 95% binomial CIs, adherence is assumed perfect and *p*_*a*_ = 0.25.

For a single pulse test, a 1-day delay can have a significant impact on the percentage of outbreaks contained, more so for shorter infection periods. For instance, for *R*_0_ = 2 (blue, thin lines), the outbreaks contained dropped from 13.4% to 0.7% for *σ* = 0.5, from 13.6% to 3.6% for *σ* = 1, and from 20.6% to 14.6% for *σ* = 2.

Similarly, the timing of the second test, *T*_2_, had a greater effect for short infection periods, again due to the separation of generations and an oscillatory behaviour similar to the one observed in Figure 1. For *σ* = 0.5 and 1, there was typically a peak in effectiveness for the second test around *T*_2_ = 4 (medium-thick lines, Panels A, B, and E) or *T*_2_ = 5 (Panel D). In Figure 2 A, a second test with *T*_2_ = 2 had no significant impact on the percentage of outbreaks contained: changing from 13.6% to just 15.4% for *R*_0_ = 2 (blue, medium-thick line). If the second test was delayed so that *T*_2_ = 4, this increased to 21.4%. Similar patterns appeared for 3 pulse tests (thick line), but with a higher effectiveness overall. In general, the wider the profile, the more likely it was that outbreaks could be contained. For instance, in the scenarios shown here and with *R*_0_ = 2, the peak percentage of outbreaks contained by 3 pulse tests for *σ* = 0.5, 1 and 2 were 25.4%, 31.2% and 56.7% respectively (blue, thick lines). This is as a consequence of the peak of the test positivity curve being fixed at 0.95, regardless of the value of *σ*, so the period of high probability of testing positive is longer for larger *σ*.

In general, as would be expected, outbreaks were less likely to be contained by any number of pulse tests for higher *R*_0_. When *R*_0_ = 3 (red lines), the infection spreads too quickly for the pulse testing to effectively contain most outbreaks. In the scenario with a wide profile (*σ* = 2) and 3 pulse tests with optimal timing ((*T*_1_, *T*_2_, *T*_3_) = (0, 2, 3)), only 24.7% of outbreaks were contained. For *σ* = 1 ((*T*_1_, *T*_2_, *T*_3_) = (0, 3, 4)) and *σ* = 0.5 ((*T*_1_, *T*_2_, *T*_3_) = (0, 4, 5)), this dropped to just 9.4% and 6.7% respectively. However, for a given infection profile and number of pulse tests, the shape of the curve shown in Figure 2 did not change substantially with *R*_0_, just shifting up and down. In particular, key features, such as the optimal timing of *T*_2_, were unchanged.

In all the scenarios shown in Figure 2, there was a significant benefit to including a second and third pulse test in terms of outbreaks contained. For example, consider the COVID-19-like profile (*σ* = 2) with *R*_0_ = 2 and *T*_1_ = 0 (panel C, blue lines): adding a second pulse test, if optimally timed, almost doubled the percentage of outbreaks contained from 20.6% up to 39.9% ((*T*_1_, *T*_2_) = (0, 2)). Introducing a third pulse test further increased it to 56.7% ((*T*_1_, *T*_2_, *T*_3_) = (0, 2, 3)).

### Comparison of Pulse Testing and Symptom-based Isolation

We now focus on the 3 pulse tests strategy with (*T*_1_, *T*_2_, *T*_3_) = (0, 1, 2) (referred to as 3-PTI from here on) and compared it to a symptom-based isolation strategy applied throughout the outbreak, with isolation starting one day after symptom onset (SI). In particular, we compare their effectiveness in terms of final outbreak size and percentage of outbreaks contained.

Figure 3 shows the final size histograms under no intervention (green), 3-PTI (red) and SI (blue), with the mean final size shown by the corresponding vertical dashed line. Each subplot corresponds to a different value of *R*_0_. We show results for *p*_*a*_ = 0.25 and *σ* = 2, corresponding to a COVID-19-like infection profile. These histograms demonstrate the qualitative difference between the two types of intervention on the outbreak final size distributions. The 3-PTI policy resulted in a binary effect where outbreaks either remained largely unaffected or drastically reduced in size. In Figure 3, Panel A (*R*_0_ = 1.5), nearly 70% of outbreaks were contained by 3-PTI, almost completely eroding the peak corresponding to large outbreaks. In Panels B and C the height of this rightmost peak in the final size distribution was reduced but the few remaining large outbreaks (i.e. the ones that are not contained) were mostly untouched. In contrast, SI reduced a few outbreaks to a minimal scale but also significantly impacted the final size of all outbreaks. For the chosen parameter values, SI narrowly outperformed 3-PTI in terms of mean final size by 0.2, 3.8 and 5.9 for *R*_0_ = 1.5, 2 and 3, respectively.

**Figure 3.**
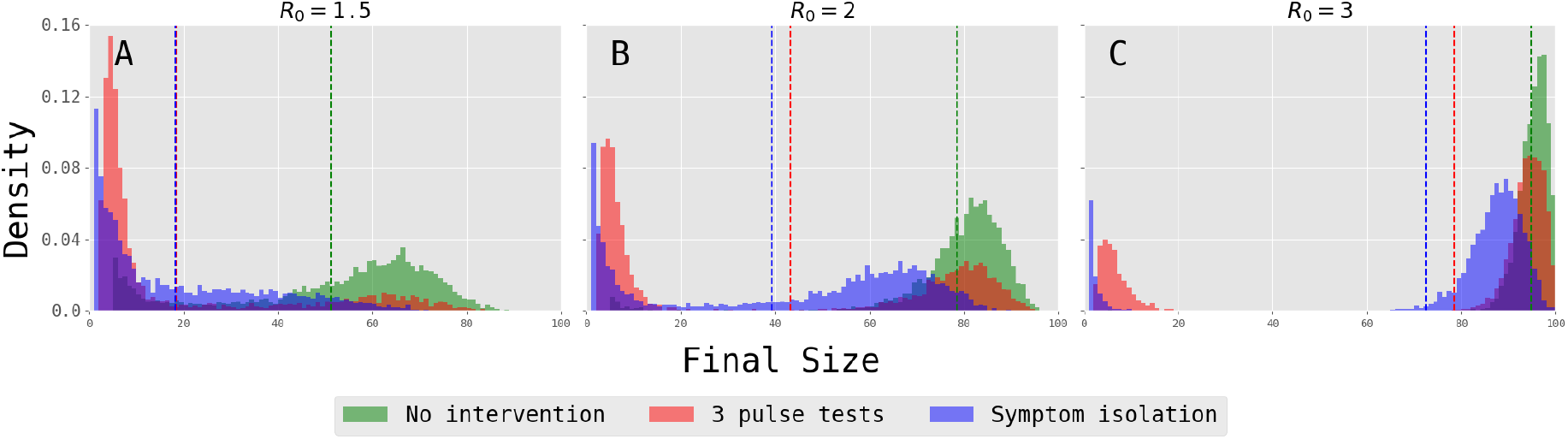
Each subplot corresponds to a different value of *R*_0_ and shows the final size distributions for three different intervention scenarios: no intervention (green), 3-PTI (red) and SI (blue). The mean of each distribution is shown by the vertical dotted line in the corresponding colour. Parameter values are *σ* = 2 and *p*_*a*_ = 0.25.

In Figure 4 we considered the effects of adherence on the effectiveness of 3-PTI (red), SI (blue), and the combination of the two (violet), in terms of percentage of outbreaks contained (solid lines). To offer a measure of the resources utilised under each intervention, we also compared the mean total number of people who are isolated throughout the outbreak (dashed lines). Each subplot shows the results for a different value of *R*_0_ (rows) and *p*_*a*_ (columns).

**Figure 4.**
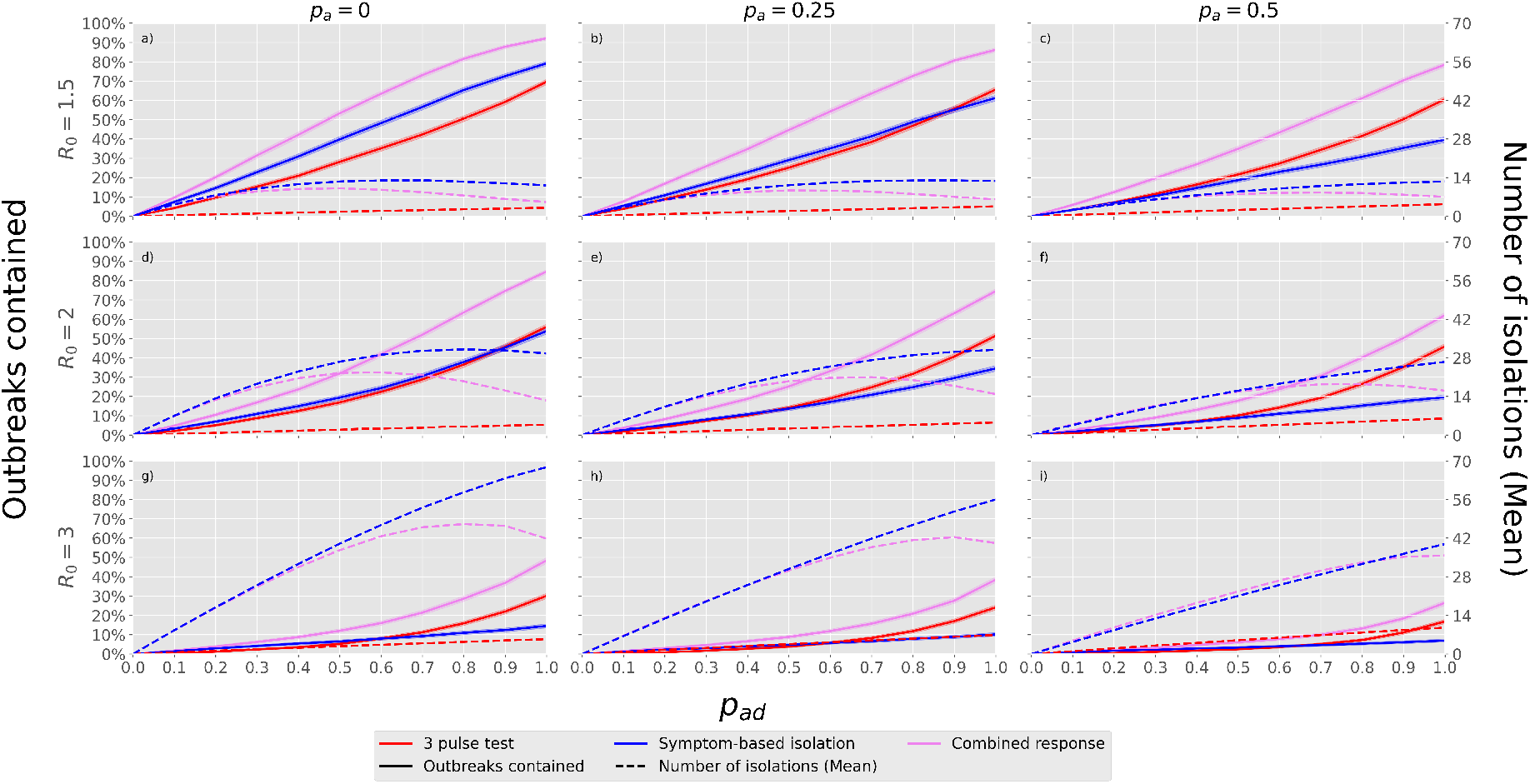
Comparison between 3-PTI (red), SI (blue) and the combination of these two (violet). Each subplot is for a different *R*_0_ (rows) and *p*_*a*_ (columns) and shows the percentage of outbreaks contained (solid) and the number of individuals isolated (dashed lines) for levels of adherence from 0% up to 100% (*p*_*ad*_, *x*-axis). Shaded areas (barely visible) show 95% CIs for the percentage of outbreaks contained. Throughout, we assume *σ* = 2.

When adherence is 100% (*p*_*ad*_ = 1), 3-PTI performed better than SI in terms of outbreaks contained in all cases except when *R*_0_ = 1.5 and *p*_*a*_ = 0. As would be expected, the gap between the effectiveness of 3-PTI and SI increased for higher *p*_*a*_: for instance, for *R*_0_ = 2, it increased from a gap of 2.2% (*p*_*a*_ = 0) to 26.3% (*p*_*a*_ = 0.5). However, the percentage of outbreaks contained by SI degraded approximately linearly with adherence compared to a faster and roughly exponential degradation for 3-PTI. This means that for lower levels of adherence, symptomatic isolation could be more effective than pulse testing. For *p*_*a*_ = 0.25 and *R*_0_ = 2, SI was slightly more effective for *p*_*ad*_ ≤ 0.4, while for *p*_*a*_ = 0 and *R*_0_ = 2 it was more effective for *p*_*ad*_ ≤ 0.8.

In all scenarios, significantly fewer people were isolated under 3-PTI compared to SI: approximately 4 to 13, 5 to 31 and 7 to 56 for *R*_0_ = 1.5, 2 and 3, respectively (*p*_*a*_ = 0.25 and *p*_*ad*_ = 1). The number of isolations for 3-PTI increased linearly with adherence. For SI, although in general higher level of adherence led to more people being isolated, the relationship was not necessarily monotonic. This can be seen in Figure 4 a) where the number of people isolated peaked at approximately 13 for *p*_*ad*_ = 0.7 compared to 11 for *p*_*ad*_ = 1, because with such high levels of effectiveness the number of isolations is overtaken by the number of potential infections that are prevented.

As well as considering the two types of intervention separately, Figure 4 shows that combining 3-PTI and SI together significantly improved the percentage of outbreaks contained. This effect was stronger for more infectious pathogens, which single interventions struggled to contain. Compared to 3-PTI, a combined response increased the percentage of outbreaks contained from 65.7% to 86.2% for *R*_0_ = 1.5, and from 24.1% to 38.5% for *R*_0_ = 3 (*p*_*a*_ = 0.25 and *p*_*ad*_ = 1).

Similarly to SI, the number of isolations needed for the combined response was non-monotonic in *p*_*ad*_, and high levels of adherence prevented enough infections to reduce the overall number of isolations needed. Compared to SI on its own, the combined response isolated approximately 7, 16 and 16 fewer individuals for *R*_0_ = 1.5, 2 and 3 (*p*_*a*_ = 0.25 and *p*_*ad*_ = 1). However, despite this effect, more people were isolated under the combined response compared to 3-PTI alone.

## Discussion

Our study investigated the effectiveness of PTI, SI and their combination in containing an outbreak as a function of the pathogen characteristics (the infectiousness *R*_0_, the probability *p*_*a*_ of being asymptomatic, and the length of the infection period, here linked to the standard deviation *σ* of the viral load curve), as well as the timing of implementation and the adherence to the intervention. An important difference of PTI and SI is that PTI was assumed to take place soon after declaration of the outbreak and aims at stopping transmission early on; on the other hand, SI was assumed to take place throughout the duration of the outbreak, aiming to reduce the transmission of symptomatic individuals and hence reduce *R*_0_ and the final size.

### Insights

For narrow infection periods (2–4 days, *σ* = 0.5, 1), new cases are likely to be infected around the same time, and generations tend to be well separated and almost synchronised. Moreover, our assumption that infectiousness and test positivity are correlated to the same viral load curve implies that there is a narrow period (centred around the peak infectiousness) over which all cases are most likely to test positive. This created an oscillatory effect where the percentage of outbreaks contained by a single pulse test peaked approximately every generation time. As the infection and test positivity profiles become broader, this effect becomes less pronounced, and generally the earlier a pulse test was implemented, the more likely it was to contain an outbreak.

In most scenarios, outbreaks were unlikely to be contained by just one pulse test. For *R*_0_ = 2, a COVID-19-like profile (*σ* = 2), and no asymptomatic cases, approximately 20% of outbreaks were contained with an immediate pulse test (*T*_1_ = 0). Including a second pulse test significantly improved the probability of an outbreak being contained; in this scenario, it increased from 20% up to a potential 35% if the second test was implemented the day after outbreak declaration ((*T*_1_, *T*_2_) = (0, 1)). Moreover, a delay in the implementation of the first test could be made up for by including a second test: for instance, a single pulse test at day 1 after outbreak declaration had a 15% probability of containing an outbreak, but including a second test at day 2 increased it to around 28%, larger than the 20% of a single pulse test with no delay from outbreak declaration, i.e. at *T*_1_ = 0. If three pulse tests are implemented, then pulse testing can perform comparably to symptom-based isolation. An optimally timed set of 3 pulse tests contained close to ≈ 55% of outbreaks for *R*_0_ = 2, and around 70% for *R*_0_ = 1.5. Further pulse tests could be considered, but continued pulse testing will become increasingly logistically challenging and additional tests are likely to face diminishing returns.

As it is modelled in this paper, SI is equivalent to a reduction in the value of *R*_0_ for the symptomatic section of the population. This meant it significantly reduced the final size for all outbreaks, particularly for values of *R*_0_, *p*_*a*_ and *σ* for which the reproduction number with the control policy implemented (i.e. averaging the full infectivity of asymptomatic cases with the partial infectivity of symptomatic cases, interrupted at isolation) was close to the threshold value of 1. The average final size was reduced from 51.4 to 18.2, from 78.5 to 39.3 and from 94.9 to 72.5 cases for *R*_0_ = 1.5, 2 and 3 respectively (*p*_*a*_ = 0.25, *σ* = 2).

Here, when only PTI was implemented, it only had an impact in the early stages of an outbreak, with no further control implemented after the last test for the rest of the outbreak. If a pulse test detects some of the active cases and successfully isolates them, this can prevent further transmission and likely interrupt the outbreak, substantially reducing the final size. However, if not enough cases are isolated (e.g. because of low adherence to testing or false negative results, either due to the imperfect test sensitivity even at peak viral load or simply to the unfortunate events of individual testing too early in the infection period to be detected), then the outbreak is more likely to still establish itself after this initial phase of pulse testing, in which case it will continue almost unaffected, i.e. with the same *R*_0_ it would have had without intervention and only a small depletion in the initial susceptible population. On the other hand, SI has a continued effect on the outbreak and acts by reducing the transmission rate of the symptomatic fraction of cases. Hence, the final size distributions were similar to those that would result from a simple reduction in *R*_0_ and therefore all outbreaks have a significant reduction in final size.

When three pulse tests were implemented quickly following the declaration of an outbreak, they performed similarly to SI with a one day delay in terms of outbreaks contained. At 100% adherence, 3-PTI outperformed SI in all but one of the pathogen scenarios we looked at, the exception being low transmission (*R*_0_ = 1.5) and no asymptomatic cases (*p*_*a*_ = 0). Although the effectiveness of both interventions was reduced when more cases are asymptomatic, the effect was much less pronounced for 3-PTI. For example, when *R*_0_ = 2 and no cases are asymptomatic, both 3-PTI and SI individually contained approximately 55% of the outbreaks. When 50% of cases were asymptomatic, this percentage was reduced to 46% for 3-PTI, and to 19% for SI.

The mean number of individuals isolated under a PTI intervention was much lower than for SI in all cases, reducing the potential impact of isolation on the wellbeing of prison residents and at the same time requiring fewer resources for the isolation. This potentially enables a more normal operation of the prison regime during an outbreak, easing the increased burden on operational staff. For 3-PTI, the number of individuals isolated had a linear relationship with adherence. This is because the number of tests is predetermined and does not change with the number of people infected – it is only whether people adhere to isolation or testing that matters. On the other hand, since SI should be implemented over the whole duration of the outbreak, the number of isolations depended on the final size of the outbreak. This meant that, for higher adherence levels, SI may on average isolate fewer people as the outbreak is more successfully reduced, although this number was still substantially higher than what required by PTI.

Since SI requires no tests and little to no additional support from clinical and public health staff, prisons might already be implementing it routinely, at least to some degree. For this reason, PTI may be considered as an additional measure that can build on SI alone to improve outcomes. Combining 3-PTI and SI can further increase the number of outbreaks contained, while also reducing the number of isolations required compared to SI alone. In the best scenario (*R*_0_ = 1.5, *σ* = 2 and *p*_*a*_ = 0) and with perfect adherence, the combined interventions contained more than 90% of the outbreaks (up from 70% for 3-PTI alone and 80% for SI alone). The effectiveness was quickly eroded by high transmissibility and low adherence. In the worst COVID-19-like pathogen scenario we considered (*R*_0_ = 3, *σ* = 2, *p*_*a*_ = 0.5), the combined interventions contained just above 25% of the outbreaks when adherence was perfect. Since it further reduces the final size in comparison with SI on its own, the combined response typically resulted in fewer isolations, particularly for high adherence.

### Limitations

In this paper, we considered a scenario in which a single case of a novel pathogen is introduced into a population with no prior immunity and assuming no further introductions. We recognise this is an idealised scenario, as staff will regularly interact with both community and prison settings. Moreover, open prisons with day release are likely to experience significant mixing between the prison and community populations. These aspects will make reintroduction likely, particularly when community prevalence is high, so that other interventions, such as quarantining upon entry (termed “reverse cohorting” in the UK [38]), test screening of staff, new prisoners and visitors, or other measures and adjustments of prison regimes should be considered in combination with PTI. However, it may be practical to quarantine and pulse test a single wing in a closed prison in a way which approaches the modelled scenario.

Since we are considering the introduction of a novel pathogen, we assume that there is no prior immunity in the population. In the presence of substantial prior immunity, either due to vaccination or natural infection (e.g. the outbreak occurs at a later stage in a pandemic, when a significant number of individuals have already been previously infected), stopping an outbreak early will be less beneficial as there are fewer possible cases to be avoided. In this scenario an intense intervention such as PTI may not be worth the significant logistical outlay.

In this model we did not consider staff and residents separately, as data to inform these contact patterns was not available. Contact patterns, even amongst residents, will not abide by the homogenous mixing assumption made here, and different types of prisons will have different contact patterns. For example, in the UK prison system, some prisons restrict movement between areas, while others allow free mixing during social and exercise times. Therefore, the degree to which our assumption of homogeneous mixing holds will depend on the prison under consideration. Including heterogeneity in contact patterns for the same average number of contacts typically leads to smaller outbreaks [39] and so adding heterogeneity will impact the performance of interventions. Including a more realistic contact network that reflects the true prison structure would also allow one to compare our results with other more sophisticated interventions, such as cell-based isolation. Also, heterogeneity in infectiousness and detectability due to symptom status or other factors was not considered.

Most of our results strongly depend on our assumption that the rate of transmission and the probability of testing positive are determined by a single underlying viral load profile. Further work into the significance of this assumption and what relations between these profiles are considered realistic are necessary. Alternative choices for parametrising these profiles would have been possible, including using a different distribution (e.g. Gamma, Weibull, lognormal) or mechanistically modelling the viral load [40, 41], though the assumption made here is flexible enough for our purposes and computationally efficient.

Finally, while investigating adherence, we assumed that individuals can equally choose to adhere to either intervention. However, in the current UK prison policy, residents can refuse to test (as testing is a medical intervention), which may hamper efficacy of PTI, but cannot refuse to isolate when directed to do so. These aspects should be taken into account for policy decisions.

## Conclusion

In this paper we have formulated a stochastic time-since-infection model and conducted a modelling study to investigate pulse testing and isolation as an NPI aimed at managing an outbreak of a novel respiratory pathogen in a prison. The aim of pulse testing is to identify and isolate both symptomatic cases as well as the presymptomatic and asymptomatic cases that made SARS-CoV-2 so hard to contain. By interrupting the outbreak at its very start, before it gets out of hand, there is a possibility that long term and large scale restrictions on freedom arising from the use of isolation can be avoided, mitigating the impact on the affected population’s mental and physical health. We considered pulse testing and isolation in comparison to, **as well as in combination with**, a more standard NPI, namely symptom-based isolation. We ran simulations under various viral load profiles, *R*_0_ values, probabilities of being asymptomatic and implementation scenarios to understand how these interventions might perform under a range of hypothetical future outbreak scenarios.

Pulse testing and isolation may be an effective intervention for containing outbreaks of novel infectious diseases if implemented well. However, this would be a substantial logistical undertaking: the outbreak would need to be declared promptly on notification of the second symptomatic case, the first test would need to be performed immediately at outbreak declaration and multiple pulse tests must be repeated quickly and with high levels of uptake in order to ensure a good chance of an outbreak being contained. Staff and material resources to undertake testing would need to be rapidly deployable. Pulse testing and isolation compares well with symptom-based isolation—a more conventional intervention—particularly for large numbers of asymptomatic cases. However, if *R*_0_ is large, or good implementation is not achieved, neither intervention is effective on its own. Pulse testing and isolation may appear to be a disproportionate reaction when only a handful of cases has been detected and its usefulness might in practice be difficult to convey to prisons and residents and assess *a posteriori*, particularly given the binary nature of its outcome. However, in some scenarios it may provide an option which quickly contains outbreaks, significantly reduces the number of individuals that would need to be isolated and the overall duration of control measures compared to symptom-based isolation alone. This may reduce the need for extra shifts for prison staff and avoid disruption to daily life and important rehabilitative activities, such as educational sessions, thus alleviating the impact on the wellbeing and health of residents and staff.

## Data Availability

All data produced in the present study are available upon reasonable request to the authors

## Acknowledgements

The authors gratefully acknowledge the UK Health Security Agency (UKHSA) for funding. FS was partially supported by the Engineering and Physical Sciences Research Council via a Mathematical Sciences Small Grant (grant number UKRI170). JX, IH and LP acknowledge support from the Wellcome Trust (grant number 227438/Z/23/Z). LP additionally acknowledges the Medical Resarch Council (grant number UKRI483). FS, JX, IH and LP are members of the JUNIPER partnership, which is supported by the Medical Research Council (grant number MR/X018598/1). FS is also a member of INdAM research group GNCS and UMI research group Modellistica Socio-Epidemiological. The views expressed are those of the author(s) and not necessarily those of the Department of Health or UKHSA.

## A Sellke Construction with Time-inhomogeneous Rates

The Sellke construction was proposed as a way to study the final size of stochastic models of epidemics [37]. This was initially restricted to a Markovian model, as considered by Bailey [42], in which the infectious period was assumed to be distributed exponentially. It was later extended by Ball [35] to study the final size distribution for more general epidemic models which allow for a general distribution for the infectious period. Further generalisations extended the method to models with heterogeneities in contact rates [43] and individual susceptibility [44].

In all these cases, it is assumed that during an individual’s (random) infectious period, there is a constant rate of infection, *λ*, which is instantaneously changed from 0 once an individual becomes infected and back to 0 once they have recovered. In reality the rate of infection is likely to be a function, *λ*(*τ*), of an individual’s time since infection *τ*. However, it can be shown that the final size of the epidemic is completely determined by the cumulative force of infection. Therefore, to simplify things, a constant infectious pressure is often assumed [45].

Although the final size of an uninterrupted epidemic is independent of the specific shape of the rate of infection, *λ*(*τ*), the success of the pulse test intervention considered in this manuscript is heavily dependent on the timing of the tests relative to the time of infection, and therefore on *λ*(*τ*). For this reason, we require a model and corresponding construction that allows for accurate simulation of the timings of infection. Here we detail an adapted Sellke construction which allows for the full simulation of outbreaks with time-inhomogeneous infection rates, which was used to obtain the results in this paper.

We consider an initial population of *n* susceptible individuals (indexed by *j* = 1, …, *n*) and *m* infected individuals (indexed by *i* = −(*m* − 1), …, −1, 0). Each initially susceptible individual is assigned a threshold value, *Q*_*j*_ for *j* = 1, …, *n*, which is exponentially distributed with mean 1. For convenience and without loss of generality we reorder and relabel these thresholds so that *Q*_1_ ≤ · · · ≤ *Q*_*n*_. Let *I*_*t*_ be the number of infected individuals at time *t*, with initially *I*_0_ = *m*. Because of the chosen ordering of individuals (and the infection process described below), infected individuals have indices *i, i* = −(*m* − 1), …, *I*_*t*_ − *m*. If individual *i* is infected during the outbreak, let *t*_*i*_ be their time of infection. Then, define the total cumulative infectious pressure towards a given susceptible individual at time *t* as:

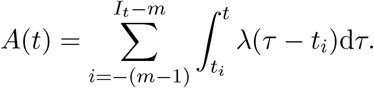

Each susceptible *j* becomes infected at the time when *A*(*t*) surpasses their threshold *Q*_*j*_, so the infection time of individual *j* is

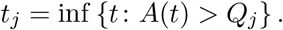

We will now verify that the Sellke construction defined above does indeed produce the transition probabilities corresponding to the model we described in this paper. Suppose *I*_*t*_ = *k* ≥ *m*, with infection times *t*_−(*m*−1)_, …, *t*_*k*−*m*_. Then, for each susceptible individual *j* (with *j > k* − *m*), we can calculate the probability that they become infected in the time interval [*t, t* + *δt*] by first calculating the complementary event:

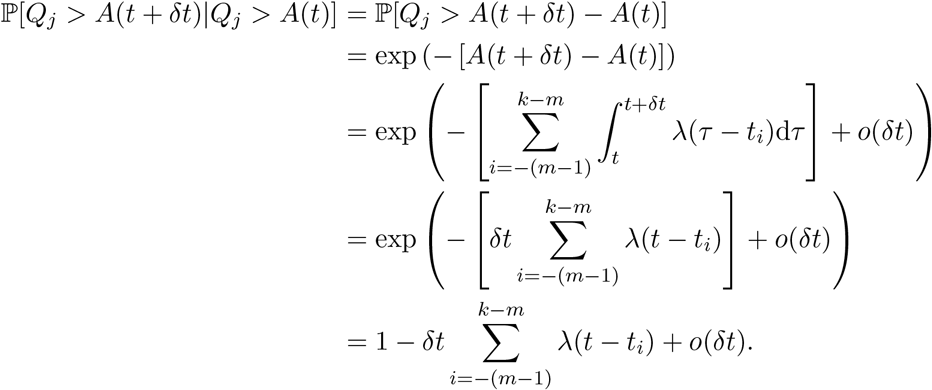

where the first equality follows from the memoryless property of the exponential distribution, the third equality results from the fact that no further infections will take place over a short enough time period, and the fourth uses the continuity of *λ* to approximate the integral. Hence, the probability that a susceptible individual is infected in the time interval [*t, t* + *δt*] is 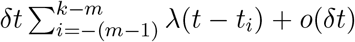. This probability is the same we would get from modelling the individual as being contacted according to the sum of each of the inhomogeneous Poisson processes from infected individuals.

